# Comparison of Inter-Joint Coordination Strategies during Activities of Daily Living with Prosthetic and Anatomical Limbs

**DOI:** 10.1101/2023.09.19.23295716

**Authors:** Christina Lee, Deanna Gates

## Abstract

While healthy individuals have redundant degrees of freedom of the joints, they coordinate their multi-joint movements such that the redundancy is effectively reduced. Achieving high inter-joint coordination may be difficult for upper limb prosthesis users due to the lack of proprioceptive feedback and limited motion of the terminal device. This study compared inter-joint coordination between prosthesis users and individuals without limb loss during different upper limb activities of daily living (ADLs). Nine unilateral prosthesis users (five males) and nine age- and sex-matched controls without limb loss completed three unilateral and three bilateral ADLs. Principal component analysis was applied to the three-dimensional motion trajectories of the trunk and arms to identify coordinative patterns. For each ADL, we quantified the cumulative variance accounted for (VAF) of the first five principal components (pcs), which was the lowest number of pcs that could achieve 90% VAF in control limb movements across all ADLs (5 < n < 9). The VAF was lower for movements involving a prosthesis compared to those completed by controls across all ADLs (p < 0.001). The pc waveforms were similar between movements involving a prosthesis and movements completed by control participants for pc1 (r > 0.78, p < 0.001). The magnitude of the relationship for pc2 and pc3 differed between ADLs, with the strongest correlation for symmetric bilateral ADLs (0.67 < r < 0.97, p < 0.001). Collectively, this study demonstrates that activities of daily living are less coordinated for prosthesis users compared to individuals without limb loss. Future work should explore how device features, such as the availability of sensory feedback or motorized wrist joints influence multi-joint coordination.

## 1. Introduction

Upper limb prostheses are prescribed to individuals with limb loss and are designed to replace the absent limb. However, most commercially-available upper limb prostheses provide limited degrees-of-freedom (DoF) of the terminal device and limited feedback (Blank, Okamura, & Kuchenbecker, 2008; Bongers, et al., 2012). These device features likely alter motor performance, as demonstrated in the form of reduced movement accuracy (Doeringer & Hogan, 1995; Lee, Gonzalez, Kang, & Gates, 2022) and quality (Bouwsema, van der Sluis, & Bongers, 2010; Engdahl & Gates, 2021) compared to anatomical movements during different upper extremity tasks.

With redundant DoF of the anatomical joints, individuals without amputation are able to use various combination of joint movement strategies to maintain their end-point performance (Cowley, Dingwell, & Gates, 2014; Cowley & Gates, 2018; Gates & Dingwell, 2008; Lee, et al., 2022; Schabowsky, Dromerick, Holley, Monroe, & Lum, 2008). While high flexibility exists, healthy individuals tend to organize multi-joint movements into a few coordinative structures (Cirstea, Mitnitski, Feldman, & Levin, 2003). This inter-joint coordination leads to several invariant features of healthy movements, such as smooth and straight movement trajectories and high end-point movement accuracy. The way movements are organized into structures can be identified using dimensionality reduction techniques such as principal component analysis (PCA) (Daffertshofer, Lamoth, Meijer, & Beek, 2004) and non-negative matrix factorization (NMF) (Rabbi, et al., 2020). Previous work has shown that the majority of movement data variance can be explained using just a few (<5) coordinative structures (Cowley & Gates, 2017; Noe, Garcia-Masso, & Paillard, 2017; Sadler, Graham, & Stevenson, 2013; St-Onge & Feldman, 2003; Tang, et al., 2019; Wang, O’Dwyer, & Halaki, 2013; Zago, et al., 2017), demonstrating a high level of coordination associated with anatomical movements.

Most upper limb prosthesis users today use a device that lack motion of the terminal device (hand) and wrist. This lack of motion likely affects the inter-joint coordination strategy used during upper extremity movements. For example, several studies have evaluated kinematic patterns in upper limb prosthesis users during different unilateral (Carey, Jason Highsmith, Maitland, & Dubey, 2008; Engdahl, Lee, & Gates, 2022; Major, Stine, Heckathorne, Fatone, & Gard, 2014) and bilateral (Carey, et al., 2008; Engdahl, et al., 2022) activities of daily living (ADLs). While there were differences across tasks, prosthesis users generally completed ADLs with increased shoulder and trunk range of motion compared to healthy individuals (Carey, et al., 2008; Engdahl, et al., 2022; Major, et al., 2014). Prosthesis users also completed ADLs with increased kinematic variability (Major, et al., 2014) and reduced movement quality, although some quality outcomes were dependent on whether the task was unilateral or bilateral (Engdahl & Gates, 2021). Previous analyses of prosthetic movements during ADLs largely focused on individual joint trajectories or characteristics of end-point performance. However, ADLs involve concurrent movement of many joints spanning across multiple planes. While existing literature has identified several performance-related movement deficits in prosthesis users, end-point performance may not accurately reflect changes in inter-joint coordination (Tomita, Rodrigues, & Levin, 2017). Therefore, analysis methods that explicitly account for multiple joint angles at once may have greater clinical utility in understanding subtle but important changes in movement coordination that occur across several joints.

Multi-joint coordination can also be affected by proprioceptive feedback (Ghez & Sainburg, 1995; Sainburg, Poizner, & Chez, 1993), or the awareness of the position of the body in space. During multi-joint movements, the most proximal joint (i.e. trunk) creates the main acceleration during a movement task, while the more distal subordinate joints (i.e. shoulder, elbow, wrist) produce their movement trajectory by taking the acceleration of the proximal joints in consideration (Dounskaia, 2005). Prior studies have found that individuals with reduced proprioception, such as those post-stroke (Laczko, Scheidt, Simo, & Piovesan, 2017) or with deafferented sensory neuropathy (Gordon, Ghilardi, & Ghez, 1995), exhibited movement deficits indicative of reduced inter-joint coordination. One of these studies further used simulation to demonstrate that these movement deficits, such as abnormal end-point jerk, were likely due to the inability to process multi-joint interaction torque (Laczko, et al., 2017). Prosthesis users also experience difficulty processing force interactions at the prosthetic joints with a limited sense of their prosthetic hand position. This lack of feedback in prosthesis users likely contribute to low end-point accuracy (Doeringer & Hogan, 1995; Lee, et al., 2022) and reduced movement quality during different upper extremity tasks (Cowley, Resnik, Wilken, Smurr Walters, & Gates, 2017; Engdahl & Gates, 2021; Lee, et al., 2022). How the reduction in proprioception affects multi-joint coordination strategy in upper limb prosthesis users remain unexplored.

The purpose of this study was to quantify differences in coordination strategies between prosthesis users and healthy individuals without amputation during unilateral and bilateral ADLs. We hypothesized that prosthetic movements would be less coordinated compared to anatomical movements during all ADLs, but that coordination would be most similar during symmetrical bilateral tasks. Furthermore, we hypothesized that prosthetic and anatomical movements will exhibit distinct dominant movement patterns.

## 2. Methods

### 2.1 Participants

Nine adults with unilateral transradial limb loss participated (Table 1). All participants had at least four months of experience using a body-powered and/or a myoelectric prosthesis. Nine age- and sex-matched healthy individuals without amputation (mean age: 45 ± 15 years, 5 males) also participated. Individuals with a history of serious visual, neurological, or musculoskeletal impairments other than upper limb loss were excluded from the study. All participants without amputation indicated right hand dominance on a handedness survey (Oldfield, 1971), except one, who was ambidextrous. All participants provided written informed consent to take part in this institutionally approved study.

**Table 1.** Participant characteristics.

| **Subject** | **Gender** | **Etiology** | **Absent Limb** | **Self-reported Prosthesis Use (hr/day)** | **Prosthesis Type** |
| --- | --- | --- | --- | --- | --- |
| **S01** | M | Acquired | Right | 6 | BP |
| **S02** | F | Acquired | Right | 8-10 | BP |
| **S03** | F | Congenital | Right | 2-3 | MYO |
| **S04** | F | Congenital | Right | 3-4 | MYO |
| **S05** | F | Congenital | Left | 3-8 | MYO |
| **S06** | M | Acquired | Right | >10 | BP |
| **S07** | M | Acquired | Right | 10 | BP |
| **S08** | M | Acquired | Left | 14 | BP |
| **S09** | M | Acquired | Right | 4 | BP |

\*BP – body-powered, MYO - myoelectric

### 2.2 Experimental protocol

Participants completed six activities of daily living (ADLs) using their intact/dominant and prosthetic/non-dominant hands. Details of each ADL can be found in (Engdahl & Gates, 2021; Engdahl, et al., 2022). Briefly, the ADLs required participants to move objects from a lower to a higher shelf (BASKET, BOX, CAN), place small objects into specific target areas (PILL, PIN), or apply deodorant (DEO). Three tasks were unilateral (CAN, PILL, PIN), two were symmetric bilateral (BASKET, BOX), and one was asymmetrical bilateral (DEO) tasks. Each ADL was performed five times at a self-selected pace. During all trials, we tracked the three-dimensional motions of the trunk, upper arms, forearms, and hands at 120 Hz using a 19-camera motion capture system (Motion Analysis, Santa Rosa, CA) and 22 reflective markers.

### 2.3 Data Analysis

Visual 3D (C-motion, Germantown, MA) filtered marker position data using a 4th-order low-pass Butterworth filter with a 10 Hz cut-off frequency. A 7-segment model was created using the joint centers and local coordinate systems defined in (Gates, Walters, Cowley, Wilken, & Resnik, 2016). Three dimensional joint angles of the trunk, shoulder, elbow, and wrist were calculated based on recommendations from the International Society of Biomechanics (Wu, et al., 2005). ADLs were segmented using a 5 cm/s velocity threshold for the wrist joint center (defined as the midpoint between the styloid markers). The movement start/stop time was also verified visually to exclude any adjustments prior to movement initiation (Engdahl & Gates, 2021). Joint angle waveforms were then time-normalized to 101 points per trial of each ADL (0-100% task completion).

For the unilateral ADLs, we included 11 joint angles: trunk lateral lean, trunk flexion, trunk axial rotation, humeral elevation, humeral plane of elevation, and humeral internal rotation, elbow flexion, forearm pronation, and wrist deviation, pronation, and flexion. Bilateral ADLs included joint angles from both limbs for a total of 19 joint angles. Prosthesis users who participated in this study did not have a motorized wrist, so the wrist joint range of motion for these individuals were set to zero degrees. Data from the first and last 10% of the movement were excluded as the shoulder angles were unreliable due to gimbal lock of the shoulder that occurs in neutral positions (near the start and stop of each trial) (Engdahl, et al., 2022; Gates, et al., 2016).

We applied principal component analysis (PCA) to find the structure of the variance in the joint angle data. PCA is a dimensionality reduction technique that outputs a new set of uncorrelated variables, which are a linear combination of the weighted original variables (Daffertshofer, et al., 2004; Deluzio & Astephen, 2007). These uncorrelated variables are referred to as principal components (pc). Specifically, we obtained the eigenvectors and eigenvalues from the covariance matrix of the original data to obtain the direction and magnitude of the pcs. The eigenvector that corresponds to the greatest eigenvalue that explains the greatest amount of variance and is considered the first principal component, or pc1. The subsequent pcs are ranked in the descending amount of variance explained. For a data set with k number of variables, there are k pcs. Typically, the number of pcs required to sufficiently explain the majority of variance in the original data set is less than k, hence effectively reducing the unique number of variables needed to describe the data.

The input features for PCA (joint angle time series) were grouped depending on the ADL. Specifically, unilateral ADL (CAN, PILL, PIN) time series were grouped by movements completed with control limb (left and right combined), prosthetic limb, and intact limb of prosthesis users. Symmetrical bilateral ADL (BOX, BASKET) time series were grouped by movements completed by control participants and prosthesis users. For asymmetrical bilateral ADLs (DEO), limb type grouping was determined by the ipsilateral limb of the object (deodorant) such that prosthetic limb group referred to trials where the prosthetic limb was used to hold and apply the deodorant. We first combined time-normalized movement trajectories for each individual (for symmetrical bilateral ADLs) or limb (for unilateral and asymmetrical bilateral ADLs) in each ADL. Within each individual/limb, we demeaned and normalized each combined joint trajectories by the range of motion.

In order to compare differences between groups, we first calculated a common set of weighting coefficients for each ADL. Movement trajectories from all control participants were concatenated for each ADL. The common weighting coefficients obtained from this concatenation were then used to calculate the pcs for each individual or limb. Across all ADLs, 5-9 pcs accounted for 90% of the variance in movements completed by control participants. Therefore, we compared the cumulative variance accounted for (VAF) by the first five pcs for each ADL per group as a measure of inter-joint coordination.

### 2.4 Statistical Analyses

For each ADL, we tested for differences in the cumulative VAF between either limb type (control, intact, prosthetic limb for unilateral and asymmetrical bilateral ADLs) or participant type (control participants and prosthesis users for symmetrical bilateral ADLs) using generalized estimating equations where group was a fixed categorical factor. Significant main effects were further explored using post-hoc pairwise comparison of estimated marginal means with a Sidak correction. All statistical analyses were performed using SPSS (version 24, IBM Corp, Armonk, NY) with α = 0.05. We assessed the similarity in average group waveforms of pc1, pc2, and pc3 for each ADL by examining the correlation coefficient in MATLAB (Natik, Massachusetts). Correlation coefficient magnitudes were considered “weak” (0 < r < 0.3), “moderate” (0.3 < r < 0.6), or “strong” (r > 0.7) (Akoglu, 2018).

## 3. Results

### 3.1 Summary of Included Data

P01 and P08 completed tasks with both a myoelectric and body-powered prosthesis. We only included data from their body-powered prosthesis as they reported wearing them more often. P01, P02 and their matched controls did not complete BOX, as the task was included in the protocol after their completion of data collection. P02 did not complete CAN and DEO due to insufficient grasp aperture of her terminal device and P03 did not complete BASKET due to pain. Movement data for P07’s matched control during BASKET was not included in the analysis due to lack of participant compliance.

### 3.2 Principal Component Weighting Coefficients

The weighting coefficient of pc1 described the dominant movement pattern required by the task (Figure 1-3; Appendix A). For example, during CAN when the participants moved a can from a low shelf to a high shelf, the dominant contributions were from humeral elevation and wrist deviation (Figure 1A, B). During BOX (symmetrical bilateral ADL), there was symmetrical loading between the left and right shoulder joint in pc1 (Figure 2A, B). In contrast, there was no symmetry between the ipsilateral (limb holding the deodorant) and the contralateral limb during DEO. Increased weighting from the contralateral shoulder represented raising the arm as individuals reached towards the armpit, which was complemented by contributions of ipsilateral elbow flexion as the participant grabbed the deodorant and brought it to the contralateral arm (Figure 3-A, B). Ipsilateral wrist contribution in pc1 likely described the swiping of the deodorant on the contralateral armpit.

**Figure 1.**
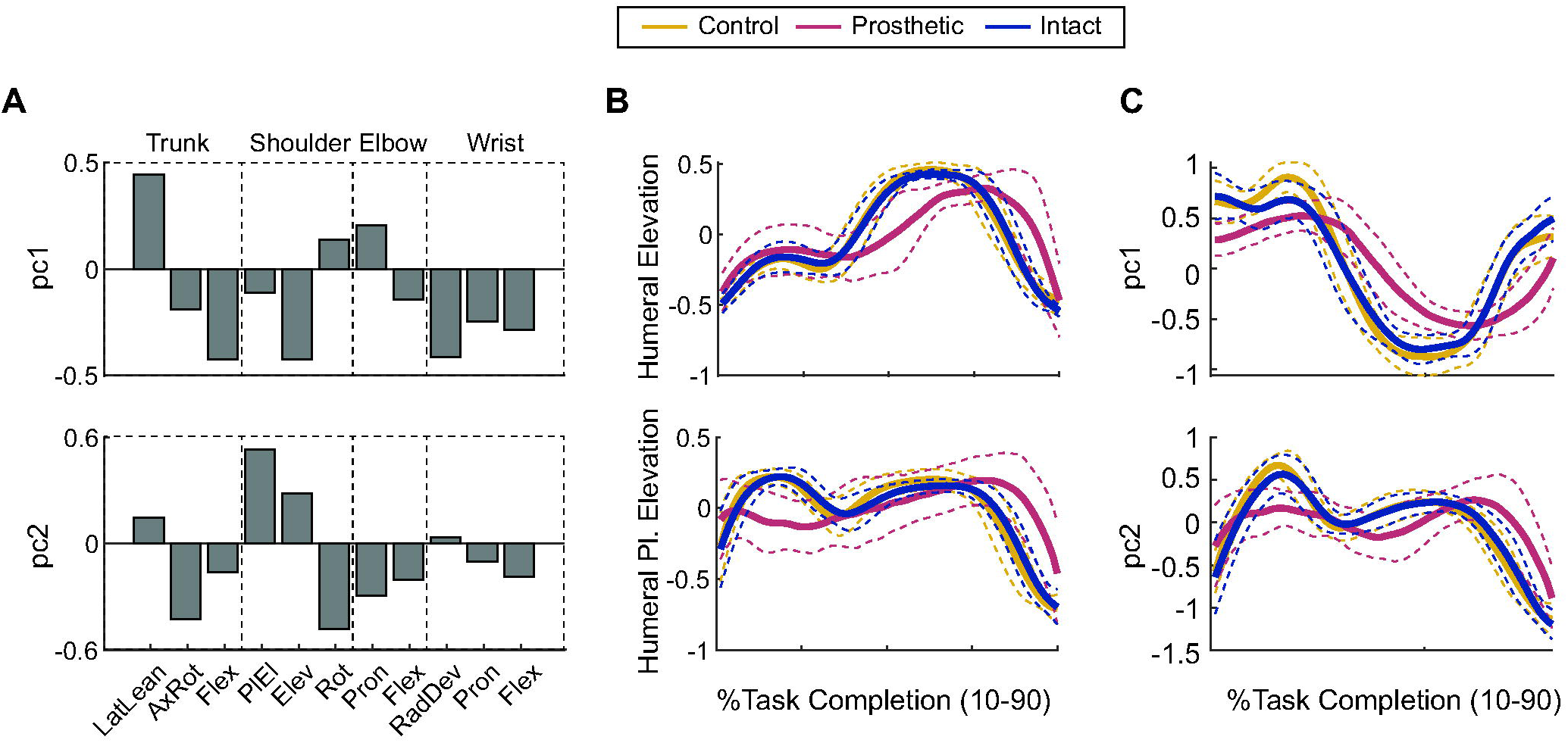
Results of CAN. (A) Common set of weighting coefficients of pc1 and pc2 during CAN, a representative unilateral ADL. The features are organized by segments – trunk, shoulder, elbow, and wrist. For the trunk, movement features were lateral lean (LatLean), axial rotation (AxRot), and flexion (Flex). For the shoulder, movement features were humeral plane of elevation (PlEl), humeral elevation (Elev), and humeral internal rotation (Rot). For the elbow, movement features were forearm pronation (Pron) and flexion (Flex). For the wrist, movement features were deviation (RadDev), pronation (Pron), and flexion (Flex). (B) Normalized average trajectory of select features (shoulder elevation for pc1 and humeral plane of elevation for pc2) that had significant weighting contributions and (C) average first and second principal component waveforms (yellow: control limb; blue: intact limb of prosthesis users; red: prosthetic limb). Standard deviations are represented as dashed lines in the same color.

**Figure 2.**
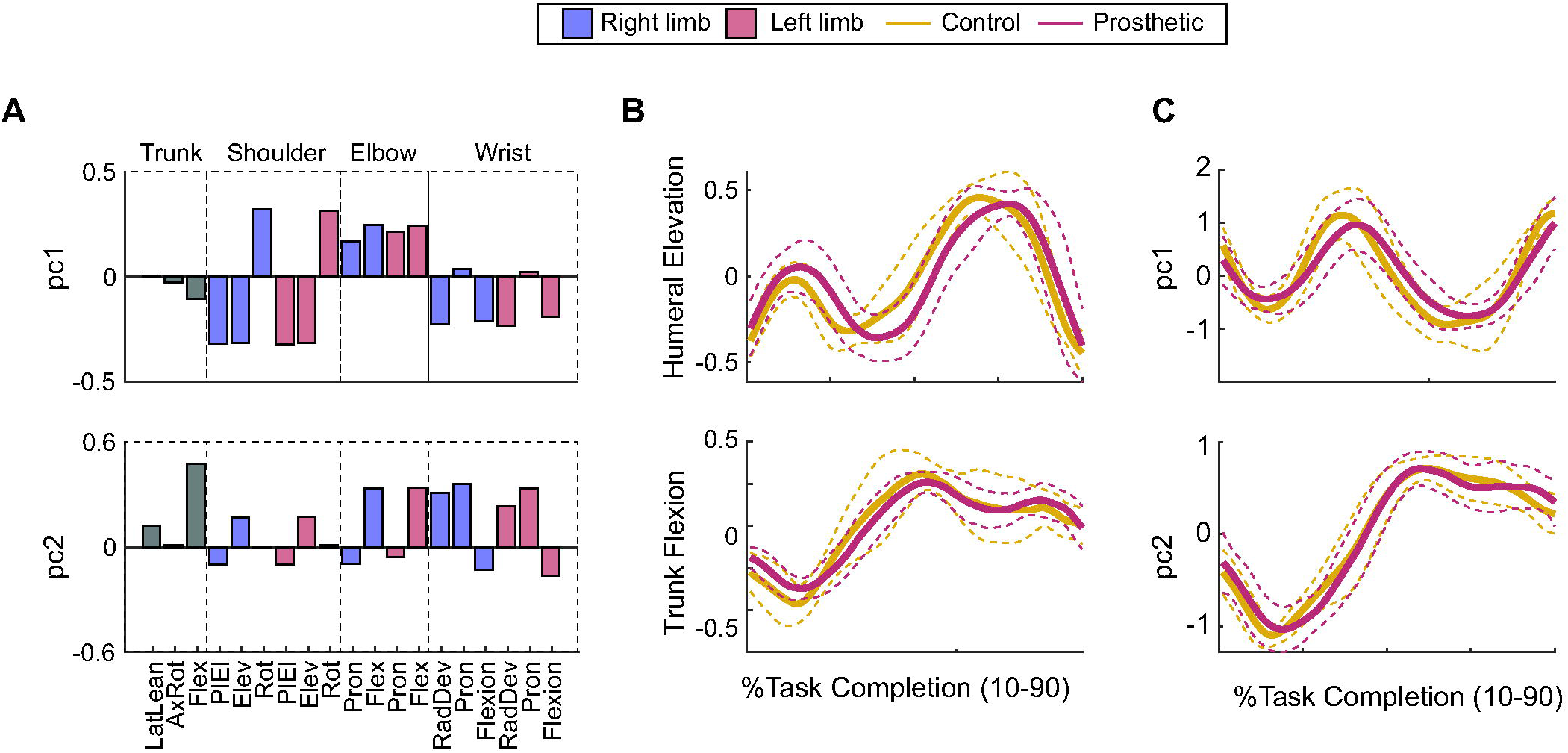
Results of BOX. (A) Common set of weighting coefficients of pc1 and pc2 during BOX, a representative symmetrical bilateral ADL. The features are organized by segments – trunk, shoulder, elbow, and wrist. For the trunk, movement features were lateral lean (LatLean), axial rotation (AxRot), and flexion (Flex). For the shoulder, movement features were humeral plane of elevation (PlEl), humeral elevation (Elev), and humeral internal rotation (Rot). For the elbow, movement features were forearm pronation (Pron) and flexion (Flex). For the wrist, movement features were deviation (RadDev), pronation (Pron), and flexion (Flex). Right and left limbs were represented as purple and red bars, respectively. (B) Normalized average trajectory of select features (shoulder elevation for pc1 and trunk flexion for pc2) that had significant weighting contributions and (C) average first and second principal component waveforms (yellow: participants; red: prosthesis users). Standard deviations are represented as dashed lines in the same color.

**Figure 3.**
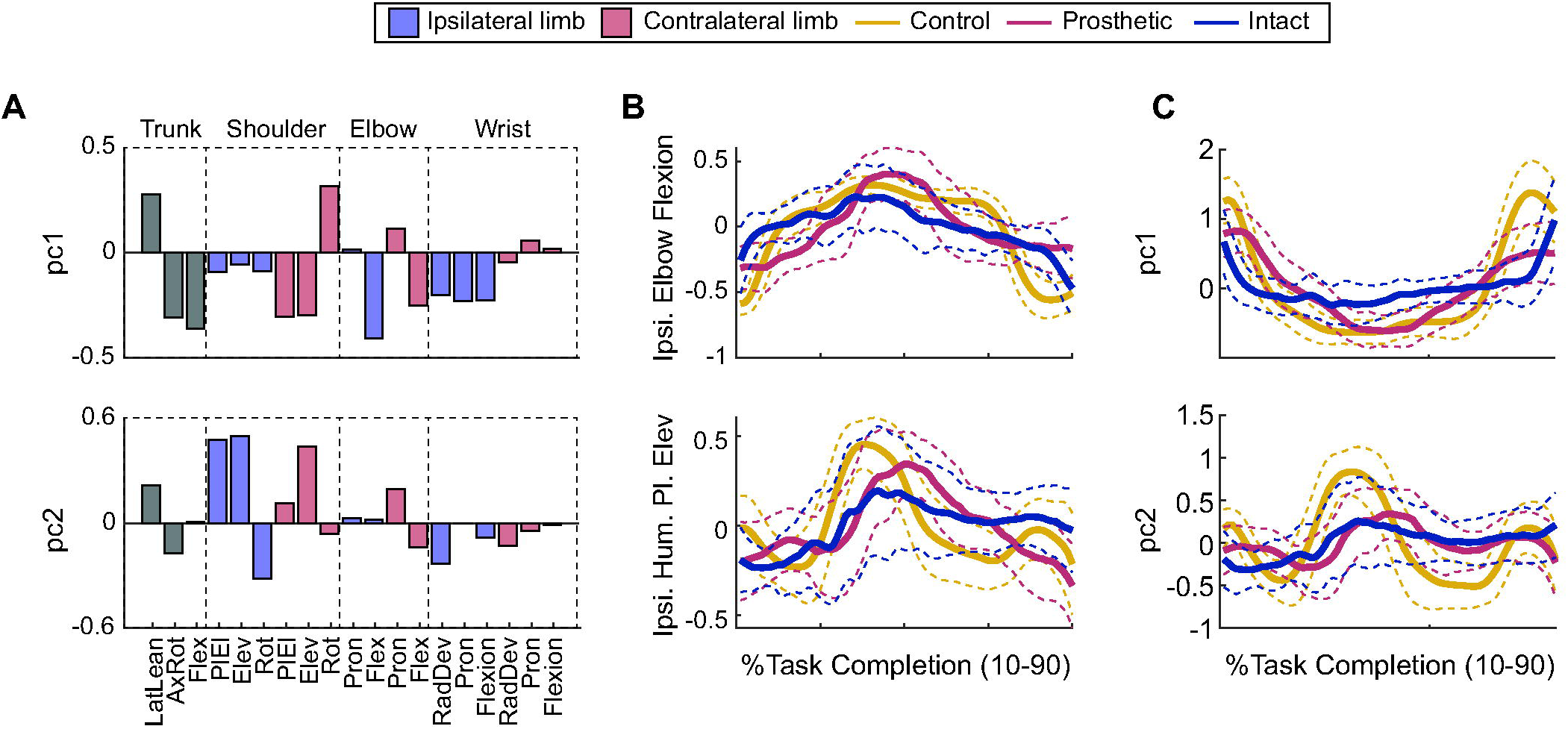
Results of DEO. (A) Common set of weighting coefficients of pc1 and pc2 during DEO, a representative asymmetrical bilateral ADL. The features are organized by segments – trunk, shoulder, elbow, and wrist. For the trunk, movement features were lateral lean (LatLean), axial rotation (AxRot), and flexion (Flex). For the shoulder, movement features were humeral plane of elevation (PlEl), humeral elevation (Elev), and humeral internal rotation (Rot). For the elbow, movement features were forearm pronation (Pron) and flexion (Flex). For the wrist, movement features were deviation (RadDev), pronation (Pron), and flexion (Flex). Ipsilateral and contralateral limbs were represented as purple and red bars, respectively. (B) Normalized average trajectory of select features (ipsilateral elbow flexion for pc1 and ipsilateral humeral plane of elevation for pc2) that had significant weighting contributions and (C) average first and second principal component waveforms (yellow: control limb; blue: intact limb of prosthesis users; red: prosthetic limb). Standard deviations are represented as dashed lines in the same color. Ipsilateral limb represents the limb holding the object (deodorant).

### 3.3 Inter-joint Coordination

The VAF of each pc differed between movements involving a prosthesis and movements completed by control participants (Figure 4). While pc1 had the greatest VAF for all groups, subsequent pcs were more variable. Often, subsequent pcs had greater VAF than the prior pcs in movements involving a prosthesis. On average, across all ADLs, pc1 and pc2 accounted for 42.1% (range 33.2-49.1%) and 18.9% (range 14.6-23.9%) of the variance in movements completed by control participants, respectively (Figure 4). For movements involving a prosthesis, pc1 and pc2 accounted for 27.3% (range 22.2-34.7%) and 12.8% (range 7.3-20.3%) of the variance, respectively. During unilateral and asymmetrical bilateral ADLs, pc1 and pc2 accounted for 33.5% (range 14.0-41.3%) and 17.0% (range 12.9-24.6%) of the variance of intact limb movements of prosthesis users, respectively.

**Figure 4.**
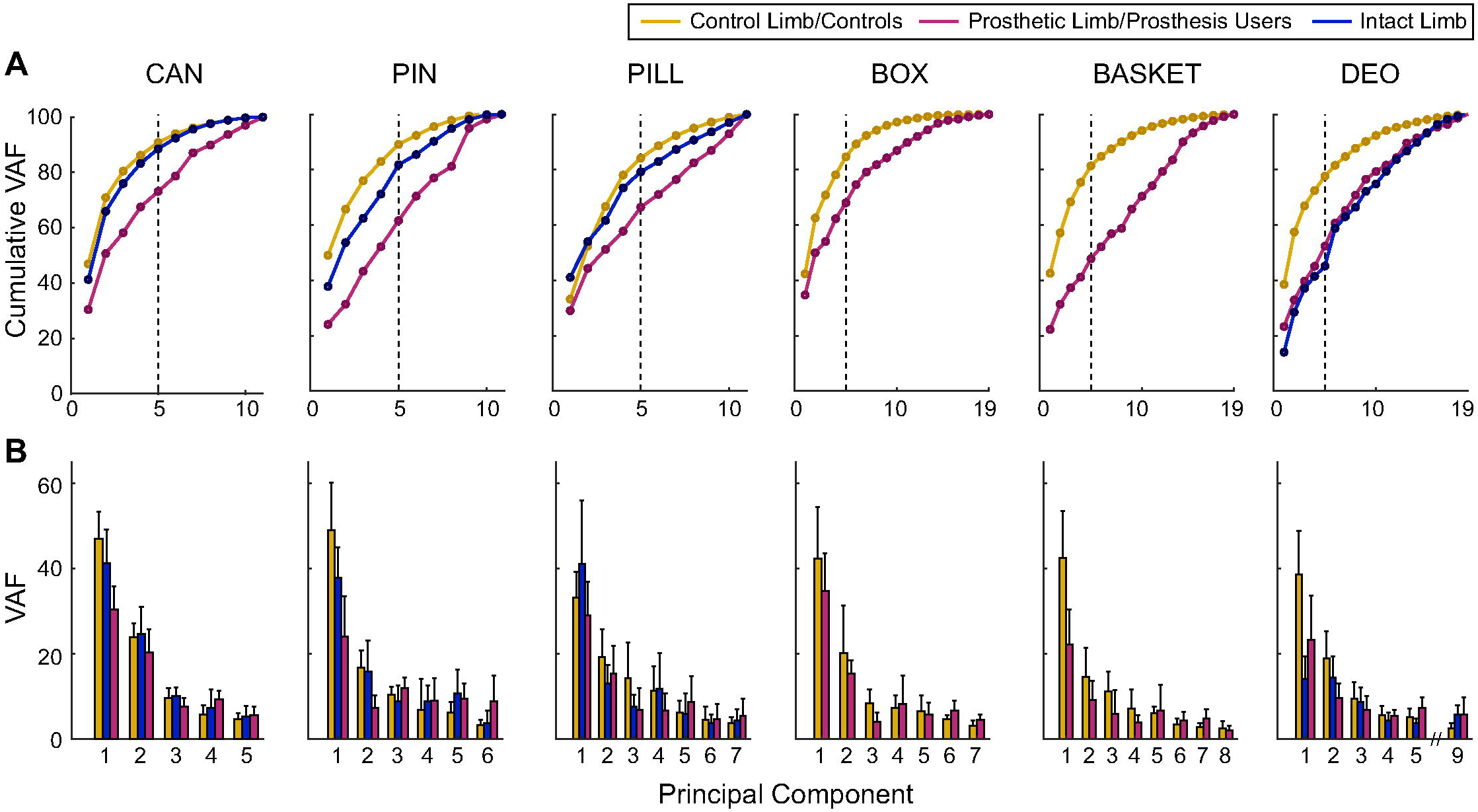
(A) Cumulative variance account for (VAF) of each principal component (pc) of each ADL for each group (yellow: control limb or control participants; red: prosthetic limb or prosthesis users; blue: intact limb of prosthesis users). Dashed line marks cumulative VAF with the first five pcs. (B) VAF of each pc of each group up to the number of pc required to explain above 90% variance of the control group (range: 5-9). The pcs were organized in a descending order such that pc1 had the most VAF for control limb or control participants. Error bar illustrates within-group variability (standard deviation).

Across all ADLs, there was a significant main effect of group in the cumulative VAF by the first five pcs (p < 0.001). Movements involving a prosthesis and intact limb movements of prosthesis users had an average cumulative VAF of 61.5% and 73.7%, respectively. For all ADLs, the cumulative VAF with the first five pcs was lower in movements involving a prosthesis compared to movements completed by control participants (p < 0.001). Intact limb movements of prosthesis users had a similar cumulative VAF during two unilateral movements (CAN and PILL; p > 0.229), but differed during another (PIN; p = 0.007). In that task, the cumulative VAF was lower for intact limb movements of prosthesis users compared to movements completed by control participants. Cumulative VAF during unilateral movements were significantly lower during prosthetic movements than intact limb movements in prosthesis users (p < 0.002). During the asymmetric bilateral task (DEO), there were no differences in the cumulative VAF between trials where the prosthesis was used to hold the deodorant (grouped as prosthetic limb) and those when the prosthesis was used to uncap the deodorant (grouped as intact limb) (p = 0.125), and both had a lower cumulative VAF compared to controls (p < 0.001).

### 3.4 Shape of Principal Component Waveforms

The shape of the first few pc waveforms were similar between movements completed by control participants and movements involving a prosthesis with an average correlation coefficient of 0.88 and 0.67 for pc1 and pc2, respectively (p < 0.001; Figure 1C, 2C, 3C; Appendix B). The third pc (pc3) tended to be less similar between groups with an average correlation coefficient magnitude of 0.38 (0.001 ≤ p < 0.68). The correlations in pc waveforms between groups were task dependent. Prosthesis users and control participants had more similar waveforms during symmetrical bilateral ADLs (average r= 0.92, 0.86, and 0.75 for pc1, pc2, and pc3, respectively) than unilateral or asymmetrical bilateral ADLs (average r=0.86, 0.57, and 0.19 for pc1, pc2, and pc3, respectively). The shape of pc1, pc2, and pc3 for the unilateral ADLs were similar for intact limb movements of prosthesis users and control limb movements (average r=0.86; p<0.001). During the asymmetrical bilateral ADL (DEO), the correlation of pc1 between control limb and intact limb was the weakest (r = 0.66, p < 0.001) out of all ADLs.

## 4. Discussion

In this study, we compared inter-joint coordination between prosthetic and anatomical movements during a series of unilateral and bilateral ADLs. For all ADLs, the cumulative variance accounted for (VAF) by the first five principal components (pcs) was lower for movements involving a prosthesis compared to movements completed by control participants. The first principal components (pc1) of movements involving a prosthesis were similar in shape as movements completed by control participants (r ≥ 0.78; Figure 1C, 2C, 3C), though they explained less variance (Figure 4). The waveforms of the subsequent pcs were less similar between groups (0.05 < |r| < 0.86) during unilateral and asymmetrical bilateral ADLs.

Comparing coordination strategies between prosthesis users and healthy individuals is difficult due to the lack of motion of the prosthetic wrist joint. To enable a more direct comparison, we chose to use a common set of weighting coefficients obtained from movements completed by healthy individuals. Using this approach, each data set has the same number of input features, enabling a more direct comparison of the coordination strategy of prosthetic users to the benchmark of control participants. In prosthesis users, the VAF of the subsequent pcs did not always decrease, which is contrary to what would be typical if the weighting coefficients were established based on prosthetic movements. The likely reason for this difference is that the weighting coefficients obtained from movements of control participants had substantial weighting on wrist motions (Figure 1-3A). While the VAF of subsequent pcs differed, the VAF of pc1 was always the greatest in all groups. A typical interpretation of pc1 is that it represents the requirements of the task (Burns, Patel, Florescu, Pochiraju, & Vinjamuri, 2017; Cowley & Gates, 2017; Zago, et al., 2017). As such, we may expect that there would be the greatest similarity between groups in pc1. As expected, pc1 waveforms had the highest correlation between movements involving a prosthesis and movements completed by control participants. However, the VAF by pc1 was lower for movements involving a prosthesis than movements completed by control participants. In subsequent pcs, the shape of the waveforms and the relative VAF were more distinct between groups, suggesting that those pcs were more dependent on limb type.

The cumulative VAF in movements completed by control participants varied for different ADLs. The average number of pcs needed to explain 90% of the variance was seven, which was greater than that reported in previous studies of upper limb movements (Cowley & Gates, 2017; Deluzio & Astephen, 2007; Noe, et al., 2017; Sadler, et al., 2013; St-Onge & Feldman, 2003; Tang, et al., 2019; Wang, et al., 2013; Zago, et al., 2017). One possible reason behind this difference in the number of principal components, or functional coordinative units, is the complexity of the tasks involved. The ADLs in this study were multi-planar and complex relative to the cyclical (Cowley & Gates, 2017; Deluzio & Astephen, 2007; St-Onge & Feldman, 2003; Zago, et al., 2017) or single-planar (Noe, et al., 2017; Sadler, et al., 2013; Tang, et al., 2019) characteristics of the tasks used in other studies. Other studies that evaluated the level of multi-joint movement organization in healthy individuals during various unstructured activities reported as many as eight principal components to explain above 80-90% of the data variance (Burns, et al., 2017; Gloumakov, Spiers, & Dollar, 2020).

While the majority of ADLs completed by intact limb of prosthesis users were performed similarly to movements completed by control participants, prosthesis users completed PIN with reduced VAF when using their intact limb compared to control participants. For this ADL, intact limb movements were completed with reduced pc3 waveform correlations (r = 0.66; p < 0.001) and cumulative VAF (p = 0.007) compared to control limb movements. Similarly, a few previous studies have found that intact limb movements of unilateral prosthesis users were completed with abnormal trajectories (Metzger, et al., 2010) or reduced accuracy (Lee, et al., 2022). These prior studies evaluated performance during planar reaching to a spatial target, while PIN required participants to perform a multi-planar reach to place a push-pin in a circular target on a corkboard. Collectively, these studies suggest that prosthesis use influences movement control of the contralateral anatomical limb during tasks focused on spatial accuracy.

During symmetrical bilateral ADLs (BOX, BASKET), the shape of pc waveforms were more similar between prosthesis users and control participants than during unilateral or asymmetrical bilateral ADLs. Nevertheless, pc1-3 were highly correlated between groups, suggesting that the dominant movement patterns were similar. Between the two symmetrical bilateral ADLs, correlations in pc waveforms were stronger for BOX (r > 0.83) than BASKET (r > 0.67). One possible reason for this this difference is that BOX did not require actuation of the terminal device in prosthesis users while BASKET did. Moreover, BASKET was a more complex task requiring manipulation of a heavier object with height and orientation variation (picking up the object from the ground and placing it on the table placed laterally). While the waveforms were similar, prosthetic movements for symmetric bilateral tasks were still completed with reduced cumulative VAF compared to controls. Previous work with this cohort found that prosthesis users match their intact limb movement quality to that of their prosthetic limb during symmetrical tasks (Engdahl & Gates, 2019). Similarly, these participants likely adjusted their intact joint movements to align with the coordinative strategy of their prosthetic limb, resulting in a reduced level of coordination.

Prosthesis users completed the asymmetrical bilateral ADL (DEO) with reduced level of coordination compared to control participants. The correlation magnitude of pc waveforms between prosthesis users and control participants decreased during DEO compared to during symmetric bilateral ADLs, particularly for pc2 and pc3. This was true regardless of which limb was the ipsilateral limb, since the ADL required substantial interaction between the object and both limbs. The reduction in pc waveform similarity in DEO demonstrates that the lack of symmetry in bilateral ADLs presents additional challenges as prosthesis users coordinate their prosthetic limb and contralateral limb together. Likely in part due to the unique coordination challenge of asymmetrical movements, several prosthesis users who participated in this study said they resort to using a cap-less deodorant or applying a deodorant to both armpits with only their intact limb at home.

There are several limitations to consider while interpreting results of this study. For instance, there were varying levels of prosthetic experiences in a relatively small cohort. However, we attempted to establish a certain baseline of expected performance by requiring participants to have at least four months of experience with their device prior to participating and choosing common everyday tasks. However, some participants expressed that they do not use their prosthesis to complete some of the more complex ADLs in this study. Rather, they relied on their intact limb or implemented modifications to the task in their daily lives. As such, some participants completed the ADLs for the first time with their prosthesis as a part of this study or were unable to complete them. Therefore, different movement strategies may have been used by these participants compared to individuals who use their prosthesis more often. That said, the fact that some participants chose not to complete common ADLs with their prosthesis re-emphasized the need to elucidate movement challenges associated with prosthetic control during these complex tasks.

In conclusion, this study demonstrated that prosthetic movements are completed with reduced level of inter-joint coordination compared to anatomical movements during a range of ADLs. Results from this work highlighted the coordination challenges prosthesis users experience during realistic and complex activities in their daily lives. Future work that continues to explore inter-joint coordination strategy in prosthesis users with sensory feedback or motorized wrist may help identify the sources behind coordination deficits experienced by conventional prosthesis users in this study.

## Data Availability

All data produced in the present study are available upon reasonable request to the authors

## Acknowledgments

The authors would like to thank Susannah Engdahl, Kelsey Ebbs, and Michael Gonzalez for assisting with the data collection. This work was supported by the Congressionally Directed Medical Research Programs under Award No. W81XWH-16-1-0648. C. Lee was supported by the Rackham Pre-doctoral Fellowship at the University of Michigan.

