## Appendix A for "Comparison of Inter-Joint Coordination Strategies during Activities of Daily Living with Prosthetic and Anatomical Limbs"

**Appendix A. Common set of weighting coefficients of principal components, representative normalized joint trajectory, and principal component waveforms of PIN, PILL, and BASKET.**

A-1. Results of PIN. (A) Common set of weighting coefficients of pc1 and pc2 during PIN. The features are organized by segments – trunk, shoulder, elbow, and wrist. For the trunk, movement features were lateral lean (LatLean), axial rotation (AxRot), and flexion (Flex). For the shoulder, movement features were humeral plane of elevation (PlEl), humeral elevation (Elev), and humeral internal rotation (Rot). For the elbow, movement features were forearm pronation (Pron) and flexion (Flex). For the wrist, movement features were deviation (RadDev), pronation (Pron), and flexion (Flex). (B) Normalized average trajectory of select features (shoulder elevation for pc1 and trunk lateral lean for pc2) that had significant weighting contributions and (C) average first and second principal component waveforms (yellow: control limb; blue: intact limb of prosthesis users; red: prosthetic limb). Standard deviations are represented as dashed lines in the same color.

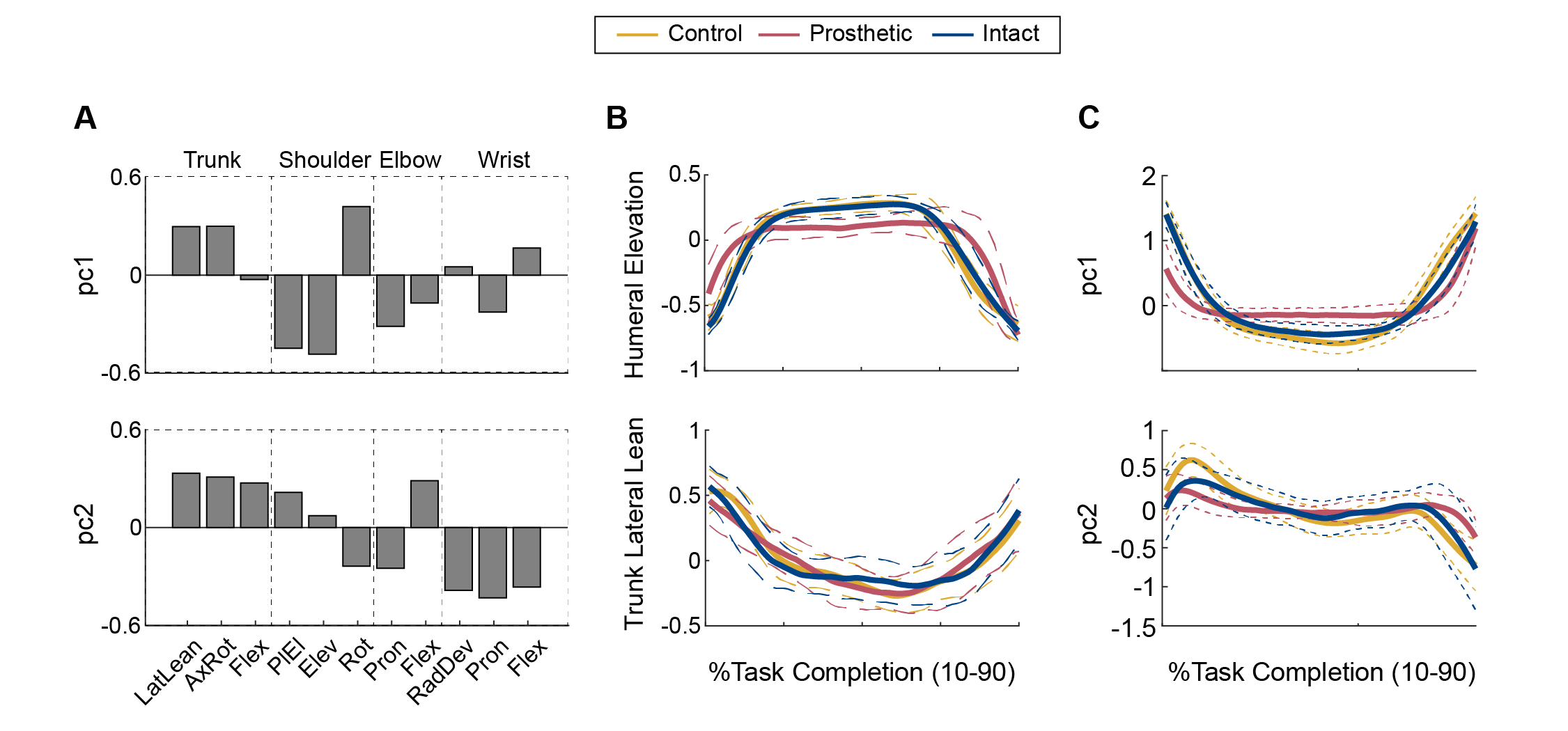

A-2. Results of PILL, (A) Common set of weighting coefficients of pc1 and pc2 during PILL. The features are organized by segments – trunk, shoulder, elbow, and wrist. For the trunk, movement features were lateral lean (LatLean), axial rotation (AxRot), and flexion (Flex). For the shoulder, movement features were humeral plane of elevation (PlEl), humeral elevation (Elev), and humeral internal rotation (Rot). For the elbow, movement features were forearm pronation (Pron) and flexion (Flex). For the wrist, movement features were deviation (RadDev), pronation (Pron), and flexion (Flex). (B) Normalized average trajectory of select features (humeral plane of elevation for pc1 and trunk flexion for pc2) that had significant weighting contributions and (C) average first and second principal component waveforms (yellow: control limb; blue: intact limb of prosthesis users; red: prosthetic limb). Standard deviations are represented as dashed lines in the same color.

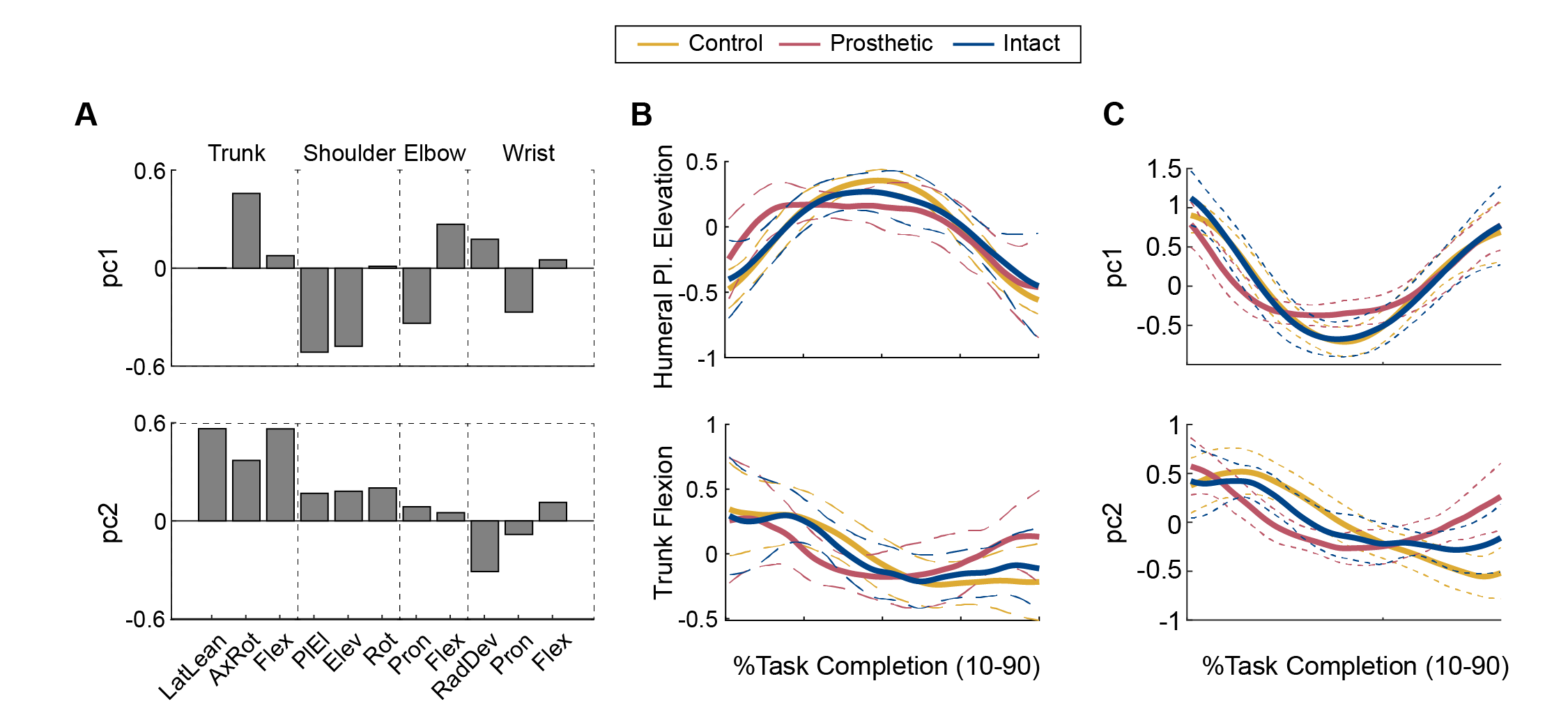

A-3. Results of BASKET. (A) Common set of weighting coefficients of pc1 and pc2 during BASKET. The features are organized by segments – trunk, shoulder, elbow, and wrist. For the trunk, movement features were lateral lean (LatLean), axial rotation (AxRot), and flexion (Flex). For the shoulder, movement features were humeral plane of elevation (PlEl), humeral elevation (Elev), and humeral internal rotation (Rot). For the elbow, movement features were forearm pronation (Pron) and flexion (Flex). For the wrist, movement features were deviation (RadDev), pronation (Pron), and flexion (Flex). Right and left limbs were represented as purple and red bars, respectively. (B) Normalized average trajectory of select features (trunk flexion for pc1 and elbow pronation for pc2) that had significant weighting contributions and (C) average first and second principal component waveforms (yellow: participants; red: prosthesis users). Standard deviations are represented as dashed lines in the same color.

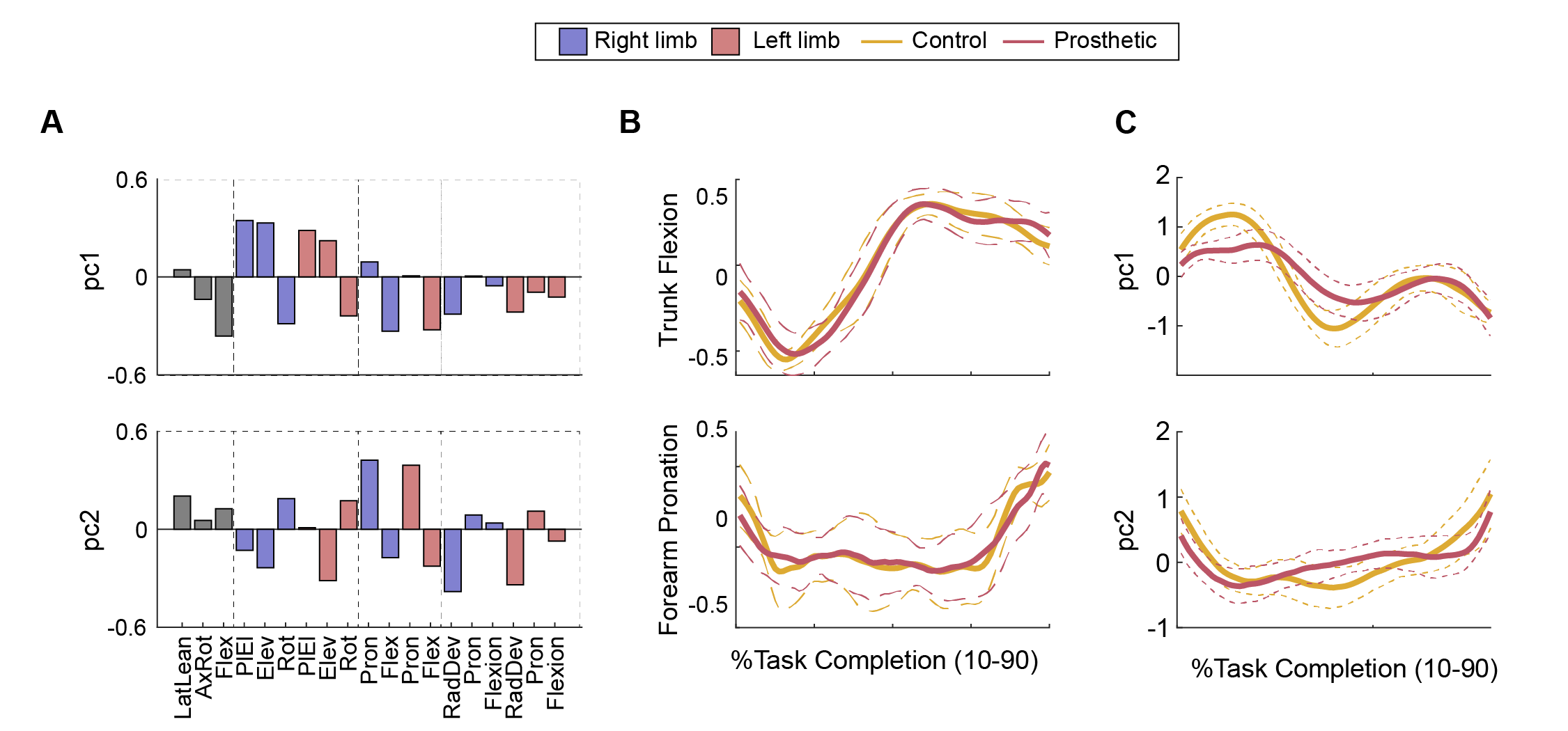
