## Appendix B for "Comparison of Inter-Joint Coordination Strategies during Activities of Daily Living with Prosthetic and Anatomical Limbs"

**Appendix B. Correlation coefficient (r) and p-value of PC 1-3 waveforms**

|  | | **Unilateral** | | | **Symmetric Bilateral** | | **Asymmetric Bilateral** |
| --- | --- | --- | --- | --- | --- | --- | --- |
| **PC1** |  | **CAN** | **PIN** | **PILL** | **BOX** | **BASKET** | **DEO** |
| **Control vs. Prosthetic** | **r** | 0.78 | 0.88 | 0.91 | 0.95 | 0.90 | 0.88 |
|  | **p** | <0.001 | <0.001 | <0.001 | <0.001 | <0.001 | <0.001 |
| **Control vs. Intact** | **r** | 0.99 | 0.99 | 0.99 |  |  | 0.79 |
|  | **p** | <0.001 | <0.001 | <0.001 |  |  | <0.001 |
| **Intact vs. Prosthetic** | **r** | 0.76 | 0.88 | 0.93 |  |  | 0.66 |
|  | **p** | <0.001 | <0.001 | <0.001 |  |  | <0.001 |

| **PC2** |  | **CAN** | **PIN** | **PILL** | **BOX** | **BASKET** | **DEO** |
| --- | --- | --- | --- | --- | --- | --- | --- |
| **Control vs. Prosthetic** | **r** | 0.67 | 0.86 | 0.37 | 0.97 | 0.75 | 0.37 |
|  | **p** | <0.001 | <0.001 | <0.001 | <0.001 | <0.001 | <0.001 |
| **Control vs. Intact** | **r** | 0.99 | 0.95 | 0.93 |  |  | 0.43 |
|  | **p** | <0.001 | <0.001 | <0.001 |  |  | <0.001 |
| **Intact vs. Prosthetic** | **r** | 0.74 | 0.86 | 0.66 |  |  | 0.63 |
|  | **p** | <0.001 | <0.001 | <0.001 |  |  | <0.001 |

| **PC3** |  | **CAN** | **PIN** | **PILL** | **BOX** | **BASKET** | **DEO** |
| --- | --- | --- | --- | --- | --- | --- | --- |
| **Control vs. Prosthetic** | **r** | 0.21 | -0.21 | -0.28 | 0.83 | 0.67 | -0.05 |
|  | **p** | 0.067 | 0.057 | 0.011 | <0.001 | <0.001 | 0.683 |
| **Control vs. Intact** | **r** | 0.90 | 0.66 | 0.93 |  |  | 0.74 |
|  | **p** | <0.001 | <0.001 | <0.001 |  |  | <0.001 |
| **Intact vs. Prosthetic** | **r** | 0.29 | 0.28 | 0.08 |  |  | 0.11 |
|  | **p** | 0.009 | 0.011 | 0.468 |  |  | 0.323 |
